# Repurposed prophylaxis strategies for COVID-19: a review

**DOI:** 10.1101/2020.05.30.20117937

**Authors:** Erwan Sallard, François-Xavier Lescure, Charles Burdet, Jérémie Guedj, Yazdan Yazdanpanah, Nathan Peiffer-Smadja

## Abstract

**Introduction:** Efficient therapeutic strategies are needed to counter the COVID-19 pandemic, caused by the SARSCoV-2 virus. In a context where specific vaccines are not yet available, the containment of the pandemic would be facilitated with efficient prophylaxis.

**Methods:** We screened several clinical trials repositories and platforms in search of the prophylactic strategies that are investigated against COVID-19 in late April 2020.

**Results:** Up to April 27, 2020, we found 68 clinical trials targeting medical workers (n = 43, 63%), patients relatives (n = 16, 24%) or individuals at risk of severe COVID-19 (n = 5, 7%). (Hydroxy)chloroquine was the most frequently evaluated treatment (n = 46, 68%), before BCG vaccine (n = 5, 7%). Sixty-one (90%) clinical trials were randomized with a median of planned inclusions of 600 (IQR 255–1515).

**Conclusion:** The investigated prophylaxis strategies cover both pre- and post-exposure prophylaxis and study numerous immune enhancers and antivirals, although most research efforts are focused on (hydroxy)chloroquine.

## INTRODUCTION

The SARS-CoV-2 is an emerging human coronavirus discovered in Wuhan, China, in December 2019. It causes the COVID-19 disease, which developed into a pandemic in early 2020: on May 24, 2020, more than 5 million persons had been infected and more than 340,000 died. In the past four months, more than 12,400 articles have been published and scientific data collected from thousands of patients have been released. This impressive research and clinical work made it possible to better understand the disease and its different phases. Numerous clinical trials are currently investigating multiple therapeutic candidates and strategies (1), including prophylaxis (2).

Prophylaxis refers to measures taken to prevent the onset of the disease. For infectious diseases it includes for example drugs aimed at blocking the infectious cycle of the pathogen or drugs that can reinforce the host immunity. There are two main categories of prophylaxis: pre-exposure prophylaxis (PrEP), where individuals who did not yet encounter the pathogen are treated, and post-exposure prophylaxis (PEP), where individuals who may have been infected (for example through contact with patients) but did not yet develop symptoms are treated. Both strategies have been extensively studied with HIV infections (3). According to these studies, PrEP with tenofovir disoproxil fumarate-emtricitabine can reduce the risk of HIV transmission by more than 90 percent in patients who are at high risk of acquiring HIV, depending on the level of adherence (4,5).

Prophylaxis is an interesting strategy for COVID-19 since it could both contain the spread of the SARSCoV-2 and prevent the development of COVID-19, especially in patients at risk of severe forms. In this review we discuss the current approaches for COVID-19 prophylaxis and the therapeutic perspectives they raise.

## METHODS

A review of currently registered clinical trials was performed to identify relevant studies. A search was conducted on April 27 on the clinicaltrials.gov repository (6), the EudraCT repository (7), the anticovid platform (8), the covid-nma platform (9) and the covid-trials platform (10), using the keywords “prophylaxis”, “PrEP” and “prevention”, except in the covid-nma platform where the keywords “healthy” and “exposed” were searched in the data file. A search on pubmed on May 14 with the key words “COVID-19 prophylaxis”, “COVID-19 prophylax*” and “COVID-19 vacc*” did not reveal any published clinical trial result.

The eligibility criteria were developed using the Patient Intervention Comparison Outcomes Study type (PICOS) framework (11).

Inclusion criteria were:

- Population: any population

- Intervention/Comparator: any antiviral agent or drug. We excluded trials evaluating therapeutic strategies whose description was not sufficient to identify a specific drug.

- Outcomes: any outcome evaluating the infection with SARS-CoV-2

- Study type: interventional clinical trial.

## RESULTS

### Number of studies

We found 309 studies on the clinicaltrials.gov repository, 32 on the EudraCT repository, 508 on the anticovid platform, 69 on the covid-nma platform and 101 on the covid-trials platform.

After eliminating the duplicates and the studies that were not testing prophylaxis (n = 951), 68 relevant clinical trials were identified (Figure 1), summarized in Table 1. Sixty-one (90%) clinical trials were randomized with a median of planned inclusions of 600 (IQR 255–1515). Most trials were focused on hydroxychloroquine (n = 46, 68%), followed by BCG vaccine (n = 5, 7%) and lopinavir/ritonavir (n = 3, 4%). The most frequently evaluated routes of administration were oral (n = 51, 75%), intradermal for vaccines (n = 6, 9%) and inhaled (n = 4, 6%). Both pre- and post-exposure prophylaxis were investigated, with a substantial number of trials on PrEP for exposed medical workers, as could be expected from the current emphasis on protecting medical workers in order to keep health systems functional through the pandemic. The complete list of trials can be found in Supplementary Table 1.

**Figure 1:**
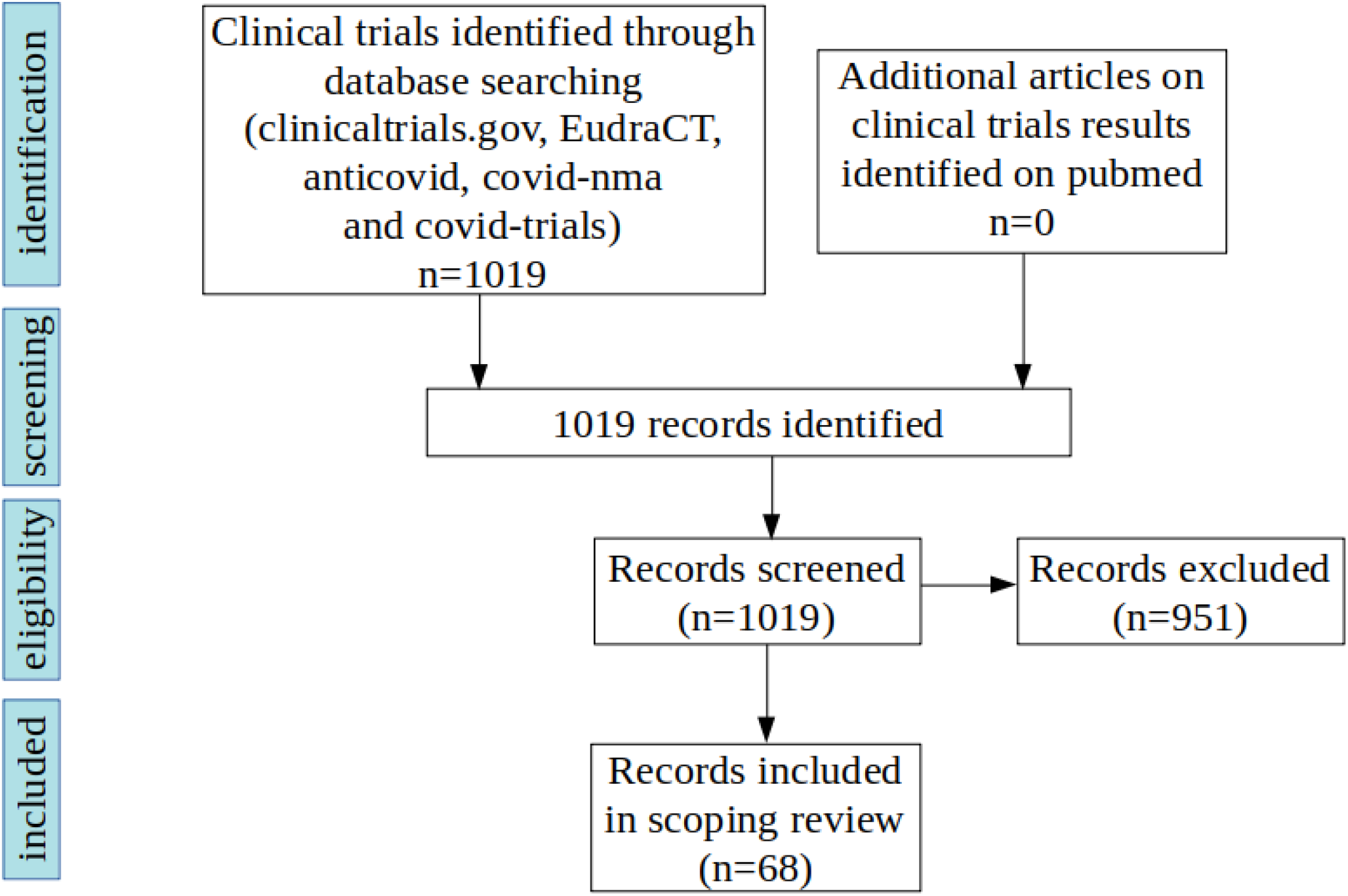
Flow chart of scientific literature search for clinical trials on prophylaxis for COVID-19

**Table 1:** Description of the clinical trials registered for the prophylaxis of COVID-19.

|  |  | N = 68 (%) |
| --- | --- | --- |
| Treatment |  |  |
|  | (Hydroxy)chloroquine* | 46 (68) |
|  | BCG vaccine | 5 (7) |
|  | Lopinavir/ritonavir | 3 (4) |
|  | Interferon | 2 (3) |
|  | Nitazoxanide | 2 (3) |
|  | Nitric oxide | 2 (3) |
|  | Azithromycin | 1 (1) |
|  | Arbidol | 1 (1) |
|  | Mycobacterium w | 1 (1) |
|  | Measles vaccines | 1 (1) |
|  | PUL-042 inhalation | 1 (1) |
|  | Convalescent serum | 1 (1) |
|  | Thiazide + calcium blocker | 1 (1) |
|  | Mesenchymal stem cells | 1 (1) |
|  | Mefloquine | 1 (1) |
|  | Melatonin | 1 (1) |
|  | Levamisole + isoprinosine | 1 (1) |
| Targeted population** |  |  |
|  | Medical workers | 43 (63) |
|  | Patients relatives | 16 (24) |
|  | At-risk individuals | 5 (7) |
|  | Others | 10 (15) |
| Administration mode |  |  |
|  | Oral | 51 (75) |
|  | Intradermal | 6 (9) |
|  | Inhaled / spray | 4 (6) |
|  | Intravenous | 2 (3) |
|  | Others | 2 (3) |
|  | Unspecified | 3 (4) |
| Study design |  |  |
|  | Randomized | 61 (90) |
|  | Non-randomized | 6 (9) |
|  | Unspecified | 1 (1) |
| Total number of planned inclusions |  |  |
|  | 200 and less | 16 (24) |
|  | 201-999 | 25 (37) |
|  | 1000 and more | 26 (38) |
|  | Unspecified | 1 (1) |

### (Hydroxy)chloroquine

Chloroquine derivatives, most notably hydroxychloroquine sulfate, inhibit coronavirus membrane fusion through an increase in endosomal pH and disrupt the glycosylation of their glycoproteins (13) They were suggested to be efficient against SARS-CoV-2 *in vitro* (12–14) and allegedly improved the disease in COVID-19 patients (17,18), although these reports are questioned and side effects are suspected (17,18). These early results and the ease to produce and administer hydroxychloroquine in high quantities may explain why it is the most investigated prophylaxis against COVID-19, with 68% (46/68) of all clinical trials analysed in this review, and involving more than 150,000 subjects in total (Supplementary Table 1). Chloroquine derivatives are administered orally at doses ranging from 400mg per week to 600mg per day, with a loading dose the first day (or occasionally the first 4 days) of 200 to 1200mg. Both pre- and post-exposure prophylaxis were represented, the former included trials conducted over 65 (IQR = 58–90) days in median, while the median duration of the latter was 5 (IQR = 5–5) days after exposure.

### Interferons

Type 1 interferons (IFN) are cytokines with pro-inflammatory and unspecific antiviral properties. Although they are produced by the organism when an infection occurs, treating COVID-19 patients with additional IFN is thought to be protective in the early phases of infection, when an acceleration in the recruitment of adaptive immunity can facilitate viral clearance (21). Therefore, type 1 interferons appear suited to prophylaxis or early disease treatment, and to immunocompromized patients (22). In late stages of the disease, an excessive immune response could be deleterious and the role of interferons is more debated. In macaques, prophylactic pegylated IFN*α*2b administered intramuscularly one on two days at 3mg/kg decreased SARS-CoV replication and lung damage (23). As a therapy, IFN*α*2b has been reported to reduce SARS-CoV-2 infection duration (24) in a small-scale, non-randomised clinical trial. Recently, IFN*α*1b was used as a prophylaxis on hundreds of health care workers, many of whom directly exposed to COVID-19 patients, and administered by nasal drops, in combination or not with thymosin-*α*1 (a putative enhancer of cellular innate immunity) (25). No COVID-19 case was reported in the individuals who received the prophylaxis. Although very promising, this result must be further confirmed, since it stems from a non-randomised clinical trial. Different modes of IFN administration are studied, notably subcutaneous pegylated IFNλ1a in a phase 2 clinical trial (NCT04344600).

### Lopinavir/ritonavir

Lopinavir/ritonavir (LPV/RTV), a protease inhibitor, was reported to improve SARS (26), although this study was criticized due to biases in patients’ assignment (27). Its safety profile is ascertained by its widespread use against HIV (28). LPV/RTV is investigated as COVID-19 post-exposure prophylaxis in the CORIPREV-LR trial (NCT04321174) and the COPEP trial (NCT04364022), during respectively 14 or 5 days following exposure to a COVID-19 patient. It is administered orally twice daily at doses of 400mg lopinavir + 100mg ritonavir in CORIPREV-LR, versus 200mg lopinavir + 50mg ritonavir in COPEP. In the trial COVIDAXIS (NCT04328285), it is used as PrEP, administered orally twice daily at doses of 200mg lopinavir + 50mg ritonavir for 2 months.

### Nitazoxanide

Nitazoxanide is a broad-spectrum antiviral that amplifies cytoplasmic RNA sensing and type 1 IFN signaling. It inhibits SARS-CoV-2 replication in vitro (14) and is tested as a PrEP for 600 elderly people in special care institutions in the trial NCT04343248, and for 800 medical workers in the trial NCT04359680. It is administered orally twice a day at doses of 600mg in both studies.

### Nitric oxide

Nitric oxide, a signaling molecule and unspecific antimicrobial, inhibits SARS-CoV replication in vitro (29) and is investigated as a PrEP for medical workers in contact with COVID-19 patients in the trial NCT04312243. It is administered twice daily through a 15 minutes long inhalation of a gas containing 160ppm of nitric oxide. In the NCT04337918 trial, it is used both as PrEP for medical workers and as post-exposure prophylaxis. Several modes of administration are investigated: gargle, nasopharyngeal irrigation and nasal spray.

### Convalescent serum

Convalescent serum intravenous administration has been proposed as a passive antibody prophylaxis or therapy against COVID-19 (30) following the hypothesis that antibodies developed by the donor, who had been infected by SARS-CoV-2 and recovered, could protect the recipient against potential infection. Convalescent serum has already been used as a therapy against MERS-CoV (31), SARS-CoV (32,33) and SARS-CoV-2 (34), and resulted in improved prognosis, but has not yet been tested as a prophylaxis. The number of recovered patients is already very high and is expected to grow further: thus, if the pool of potential donors is efficiently harnessed, large quantities of convalescent serum could be produced and convalescent plasma may become a good candidate for large-scale prophylaxis. Therefore, convalescent serum has been included in the guidelines of the Infectious Diseases Society of America (35) for both pre- and post-exposure prophylaxis. This treatment will be tested as a post-exposure prophylaxis, with 150 individuals belonging to categories highly susceptible to develop a severe disease, in a phase 2 clinical trial (NCT04323800).

However, the most relevant dose of convalescent serum has yet to be determined, and convalescent serum treatment raises the risk of antibody-dependant enhancement of infection (ADE), a process observed in a few coronaviruses (36). Consequently, investigations aiming to determine if convalescent antibodies for SARS-CoV-2 could induce ADE are warranted.

An alternative to convalescent serum prophylaxis is the use of antibody preparations, which are already being developed, but we did not find clinical trials investigating them.

### Tuberculosis or measles vaccines

The BCG tuberculosis vaccine is known to have non-specific protective effects against respiratory infections. Moreover, the geographical distribution of BCG vaccination is negatively correlated with the prevalence and mortality of COVID-19 (37,38), although the significance of this correlation is debated (39,40). 5 clinical trials of BCG vaccination on medical workers exposed to COVID-19 are being conducted (NCT04328441, NCT04327206, NCT04348370, NCT04350931, NCT04362124) and involve together 8810 subjects.

Similarly, Mycobacterium w, another tuberculosis vaccine, is tested as anti COVID-19 prophylaxis (both pre- and post-exposure) in the trial NCT04353518.

An *in silico* comparison of SARS-CoV-2 proteins with those of the measles, mumps and rubella viruses suggested that the antigens of the MMR vaccine may immunise patients against SARS-CoV-2 epitopes (41). Although this hypothesis has not yet been tested *in vitro* or *in vivo*, it prompted the launch of the NCT04357028 trial, where the MMR vaccine is used as PrEP for medical workers.

### Levamisole and isoprinosine

Levamisole is a stimulator of T helper type 1 immune response (42), used as vaccine adjuvant. Isoprinosine is also an immunostimulator and has antiviral properties whose mechanism is unclear (43). These two drugs are tested in combination in the trial NCT04360122, as a PrEP for medical workers. They are administered orally at doses of 150mg levamisole twice a week and 1g isoprinosine three times a day.

### Other treatments

Other compounds are investigated in combination with hydroxychloroquine. Umifenovir (arbidol) is a broad-spectrum antiviral approved in Russia and China which impairs viral membrane fusion (44) and displays anti SARS-CoV-2 effects. It was correlated with improvements in COVID-19 in a small-scale (16 patients in the arbidol group), non-randomised clinical trial (45). It is investigated as post-exposure prophylaxis in the trials ChiCTR2000029803 and ChiCTR2000029592. Azithromycine is an antibiotic and antiviral reported to synergize with hydroxychloroquine against COVID-19 in the controversial report of Gautret *et al*.(18). It is compared with hydroxychloroquine as PrEP for medical workers in the trials NCT04344379 and NCT04354597, with an oral administration of respectively 250mg per day and 500mg per week.

Mefloquine, an anti-malarial drug, was identified as a potent in vitro inhibitor of SARS-CoV-2 (46) and other coronaviruses (47,48), and is currently evaluated as a post-exposure prophylaxis in the 2020-001194-69 clinical trial on 200 individuals, with an oral administration of 250mg/day for a month.

In the NCT04334928 trial, both tenofovir disoproxil and emtricitabine are tested. These are nucleoside inhibitors of HIV reverse transcriptase. Their use against COVID-19 was probably prompted by the discovery that tenofovir binds SARS-CoV-2 RdRp (49), suggesting an antiviral effect. They are administered orally at respective doses of 245 and 200 mg per day to medical workers.

Bromhexine is a potent and specific inhibitor of the TMPRSS2 protease involved in SARS-CoV-2 spike protein maturation (50). It is investigated in an early phase 1 clinical trial (NCT04340349), where it is administered orally at doses of 8mg 3 times a day.

**Figure 2:**
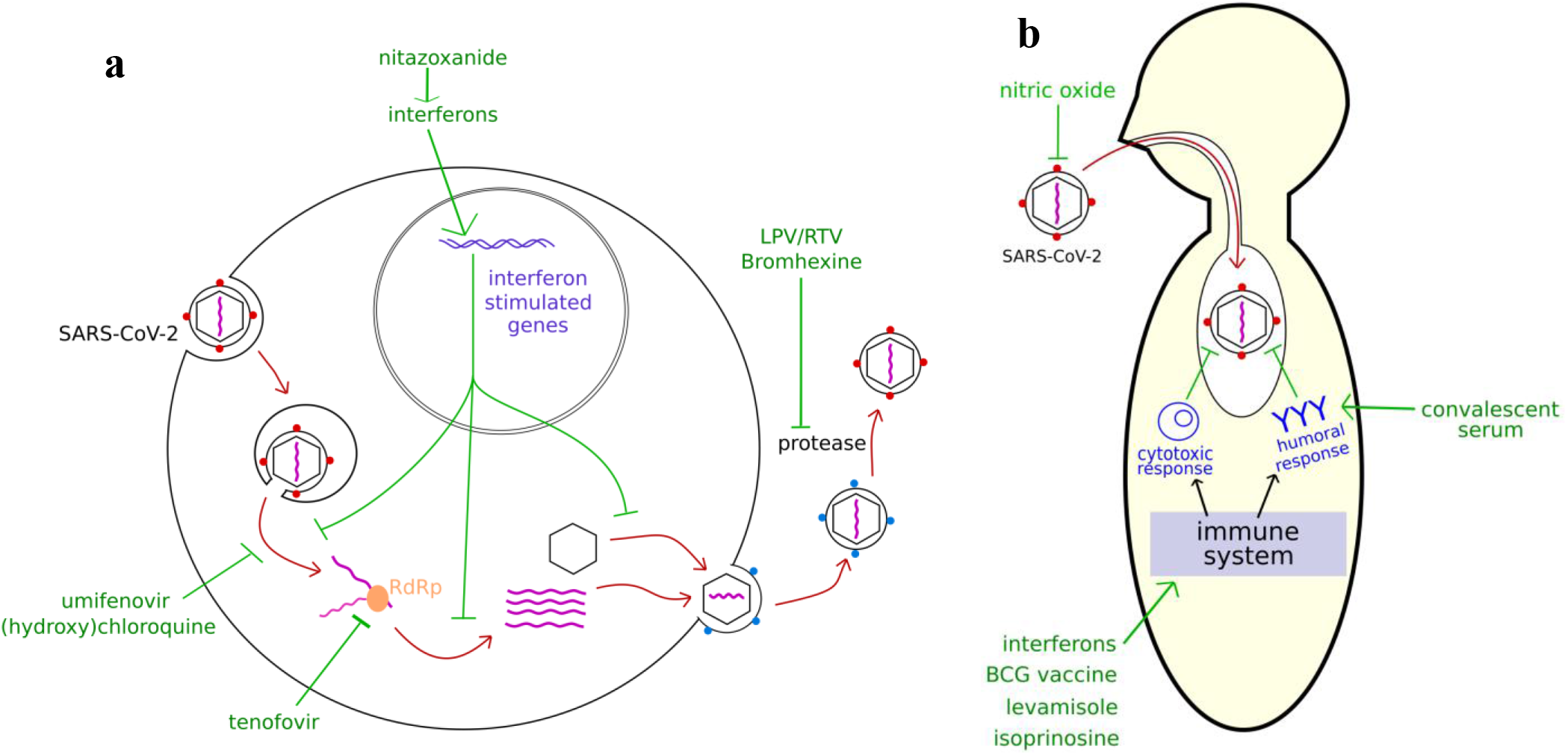
rationale for the use of the different prophylaxis strategies against COVID-19. **a**: mode of action of different antivirals in SARS-CoV-2 life cycle. **b**: mode of action of a few prophylactic strategies on COVID-19 patients

## DISCUSSION

Most of the trials studied here are randomised and include a large number of patients. Prophylaxis research efforts are mainly concentrated on (hydroxy)chloroquine although evidence in favour of this drug is currently low, a fact which has already been reported for clinical therapeutic trials (1). Numerous other antivirals potentially active on SARS-CoV-2 are investigated, but in a limited number of studies. Half of the trials on immune enhancers are testing vaccines against tuberculosis or measles, notably the BCG vaccine.

Both pre and post exposure prophylaxis are investigated. PrEP strategies targeted at-risk individuals (such as elderly or with chronic medical conditions such as obesity (12)) or, in most cases, medical workers highly exposed to infectious patients, on the protection of whom a special emphasis is put in order to keep health systems functional through the pandemic.

Most trials were focused on (hydroxy)chloroquine (68%), which explains the prevalence of orally administered treatments. Numerous trials on hydroxychloroquine or BCG vaccine are redundant because they follow identical or very similar protocols.

An important challenge with prophylactic treatments is that they must be pursued or repeated until the recipient is immunized or falls out of the priority categories, because the protective effects are short-lived: from a few hours with interferon nasal drops (25) to a few weeks with convalescent serum (30).The long duration of treatments and the fact that they are targeted on healthy individuals make it essential to propose treatments easily administered on an outpatient basis and with an excellent tolerance. Compared with therapeutic treatments, more risks are taken and less advantages are expected, which may lead to exclude treatments such as hydroxychloroquine for which side effects have been reported.

Naturally, the prophylactic strategies evaluated are centered on the early antiviral action of drugs or the stimulation of the immune system, e.g. with interferons or convalescent antibodies. The anti-inflammatory strategies that have been described elsewhere are reserved for patients with severe disease and an excessive immune response to the virus (51).

We only included trials that were registered up to April 27, 2020 but new approaches could be tested in future trials, notably antivirals that demonstrated prophylactic efficiency against coronaviruses, such as EIDD-2801 (52).

## CONCLUSION

Numerous strategies of prophylaxis against COVID-19 are currently investigated, and target different steps of the virus life cycle or the patient immune system. (Hydroxy)chloroquine is being evaluated in 68% of the registered clinical trials that we found, while numerous prophylactic strategies were investigated in a small number of trials. This discrepancy highlights the need to increase the number of treatments investigated in order to achieve an extensive cover of all promising candidate treatments against COVID-19.

## Data Availability

All data used in this article stems from registered clinical trials and is summarised in the supplementary table.

## REFERENCES

1. Belhadi D, Peiffer-Smadja N, Lescure F-X, Yazdanpanah Y, Mentré F, Laouénan C. A brief review of antiviral drugs evaluated in registered clinical trials for COVID-19. medRxiv. 2020 Mar 27;2020.03.18.20038190.

2. Fragkou PC, Belhadi D, Peiffer-Smadja N, Moschopoulos CD, Lescure F-X, Janocha H, et al. Review and methodological analysis of trials currently testing treatment and prevention options for the novel coronavirus disease (COVID-19) globally. [Internet]. Infectious Diseases (except HIV/AIDS); 2020 May [cited 2020 May 4]. Available from: http://medrxiv.org/lookup/doi/10.1101/2020.04.27.20080226

3. Krakower DS, Jain S, Mayer KH. Antiretrovirals for Primary HIV Prevention: the Current Status of Pre- and Post-exposure Prophylaxis. Curr HIV/AIDS Rep. 2015 Mar 1;12(1):127–38.

4. Molina J-M, Capitant C, Spire B, Pialoux G, Cotte L, Charreau I, et al. On-Demand Preexposure Prophylaxis in Men at High Risk for HIV-1 Infection. New England Journal of Medicine. 2015 Dec 3;373(23):2237–46.

5. Chou R, Evans C, Hoverman A, Sun C, Dana T, Bougatsos C, et al. Preexposure Prophylaxis for the Prevention of HIV Infection: Evidence Report and Systematic Review for the US Preventive Services Task Force. JAMA. 2019 Jun 11;321(22):2214–30.

6. Home – ClinicalTrials.gov [Internet]. [cited 2020 Apr 28]. Available from: https://www.clinicaltrials.gov/

7. EU clinical trials register [Internet]. [cited 2020 Apr 27]. Available from: https://www.clinicaltrialsregister.eu

8. COVID-19 trials platform by Inato [Internet]. [cited 2020 Apr 23]. Available from: https://covid.inato.com/analysis

9. elise. Covid-19 – Living NMA [Internet]. Covid-19 – Living NMA. [cited 2020 Apr 28]. Available from: https://covid-nma.com/

10. Thorlund K, Dron L, Park J, Hsu G, Forrest JI, Mills EJ. A real-time dashboard of clinical trials for COVID-19. The Lancet Digital Health [Internet]. 2020 Apr 24 [cited 2020 Apr 28]; Available from: https://www.thelancet.com/journals/landig/article/PIIS2589-7500(20)30086-8/abstract

11. Schardt C, Adams MB, Owens T, Keitz S, Fontelo P. Utilization of the PICO framework to improve searching PubMed for clinical questions. BMC Med Inform Decis Mak. 2007 Dec;7(1):16.

12. Wynants L, Van Calster B, Bonten MMJ, Collins GS, Debray TPA, De Vos M, et al. Prediction models for diagnosis and prognosis of covid-19 infection: systematic review and critical appraisal. BMJ. 2020 Apr 7;m1328.

13. Vincent MJ, Bergeron E, Benjannet S, Erickson BR, Rollin PE, Ksiazek TG, et al. Chloroquine is a potent inhibitor of SARS coronavirus infection and spread. Virol J. 2005;2(1):69.

14. Wang M, Cao R, Zhang L, Yang X, Liu J, Xu M, et al. Remdesivir and chloroquine effectively inhibit the recently emerged novel coronavirus (2019-nCoV) in vitro. Cell Res. 2020 Mar;30(3):269–71.

15. Liu J, Cao R, Xu M, Wang X, Zhang H, Hu H, et al. Hydroxychloroquine, a less toxic derivative of chloroquine, is effective in inhibiting SARS-CoV-2 infection in vitro. Cell Discov. 2020 Dec;6(1):16.

16. Yao X, Ye F, Zhang M, Cui C, Huang B, Niu P, et al. In Vitro Antiviral Activity and Projection of Optimized Dosing Design of Hydroxychloroquine for the Treatment of Severe Acute Respiratory Syndrome Coronavirus 2 (SARS-CoV-2). Clinical Infectious Diseases. 2020 Mar 9;ciaa237.

17. Gao J, Tian Z, Yang X. Breakthrough: Chloroquine phosphate has shown apparent efficacy in treatment of COVID-19 associated pneumonia in clinical studies. BioScience Trends. 2020;14.

18. Gautret P, Lagier J-C, Parola P, Hoang VT, Meddeb L, Mailhe M, et al. Hydroxychloroquine and azithromycin as a treatment of COVID-19: results of an open-label non-randomized clinical trial. International Journal of Antimicrobial Agents. 2020 Mar;105949.

19. Lane JCE, Weaver J, Kostka K, Duarte-Salles T, Abrahao MTF, Alghoul H, et al. Safety of hydroxychloroquine, alone and in combination with azithromycin, in light of rapid wide-spread use for COVID-19: a multinational, network cohort and self-controlled case series study [Internet]. Rheumatology; 2020 Apr [cited 2020 May 4]. Available from: http://medrxiv.org/lookup/doi/10.1101/2020.04.08.20054551

20. Magagnoli J, Narendran S, Pereira F, Cummings T, Hardin JW, Sutton SS, et al. Outcomes of hydroxychloroquine usage in United States veterans hospitalized with Covid-19 [Internet]. Infectious Diseases (except HIV/AIDS); 2020 Apr [cited 2020 May 4]. Available from: http://medrxiv.org/lookup/doi/10.1101/2020.04.16.20065920

21. Sallard E, Lescure F-X, Yazdanpanah Y, Mentre F, Peiffer-Smadja N. Type 1 interferons as a potential treatment against COVID-19. Antiviral Research. 2020 Jun;178:104791.

22. Hadjadj J, Yatim N, Barnabei L, Corneau A, Boussier J, Pere H, et al. Impaired type I interferon activity and exacerbated inflammatory responses in severe Covid-19 patients [Internet]. Infectious Diseases (except HIV/AIDS); 2020 Apr [cited 2020 May 4]. Available from: http://medrxiv.org/lookup/doi/10.1101/2020.04.19.20068015

23. Haagmans BL, Kuiken T, Martina BE, Fouchier RAM, Rimmelzwaan GF, van Amerongen G, et al. Pegylated interferon-α protects type 1 pneumocytes against SARS coronavirus infection in macaques. Nat Med. 2004 Mar;10(3):290–3.

24. Zhou Q, Wei X-S, Xiang X, Wang X, Wang Z-H, Chen V, et al. Interferon-a2b treatment for COVID-19 [Internet]. Infectious Diseases (except HIV/AIDS); 2020 Apr [cited 2020 May 4]. Available from: http://medrxiv.org/lookup/doi/10.1101/2020.04.06.20042580

25. Meng Z, Wang T, Li C, Chen X, Li L, Qin X, et al. An experimental trial of recombinant human interferon alpha nasal drops to prevent coronavirus disease 2019 in medical staff in an epidemic area [Internet]. Infectious Diseases (except HIV/AIDS); 2020 Apr [cited 2020 May 4]. Available from: http://medrxiv.org/lookup/doi/10.1101/2020.04.11.20061473

26. Chu CM. Role of lopinavir/ritonavir in the treatment of SARS: initial virological and clinical findings. Thorax. 2004 Mar 1;59(3):252–6.

27. Stockman LJ, Bellamy R, Garner P. SARS: Systematic Review of Treatment Effects. Low D, editor. PLoS Med. 2006 Sep 12;3(9):e343.

28. Hicks C, King M, Gulick R, White C, Eron J, Kessler H. Long-term safety and durable antiretroviral activity of lopinavir/ritonavir in treatment-naive patients: 4 year follow-up study. AIDS.2004;18:775–9.

29. Åkerström S, Mousavi-Jazi M, Klingström J, Leijon M, Lundkvist Å, Mirazimi A. Nitric Oxide Inhibits the Replication Cycle of Severe Acute Respiratory Syndrome Coronavirus. JVI. 2005 Feb 1;79(3):1966–9.

30. Casadevall A, Pirofski L. The convalescent sera option for containing COVID-19. Journal of Clinical Investigation. 2020 Mar 13;130(4):1545–8.

31. Ko J-H, Seok H, Cho SY, Ha YE, Baek JY, Kim SH, et al. Challenges of convalescent plasma infusion therapy in Middle East respiratory coronavirus infection: a single centre experience. Antivir Ther (Lond). 2018;23(7):617–22.

32. Cheng Y, Wong R, Soo YOY, Wong WS, Lee CK, Ng MHL, et al. Use of convalescent plasma therapy in SARS patients in Hong Kong. Eur J Clin Microbiol Infect Dis. 2005 Jan;24(1):44–6.

33. Yeh K-M, Chiueh T-S, Siu LK, Lin J-C, Chan PKS, Peng M-Y, et al. Experience of using convalescent plasma for severe acute respiratory syndrome among healthcare workers in a Taiwan hospital. Journal of Antimicrobial Chemotherapy. 2005 Nov 1;56(5):919–22.

34. Shen C, Wang Z, Zhao F, Yang Y, Li J, Yuan J, et al. Treatment of 5 Critically Ill Patients With COVID-19 With Convalescent Plasma. JAMA. 2020 Apr 28;323(16):1582.

35. IDSA COVID-19 guidelines [Internet]. Available from: https://www.idsociety.org/practiceguideline/covid-19-guideline-treatment-and-management/

36. Wan Y, Shang J, Sun S, Tai W, Chen J, Geng Q, et al. Molecular Mechanism for Antibody-Dependent Enhancement of Coronavirus Entry. Gallagher T, editor. J Virol. 2019 Dec 11;94(5):e02015–19, /jvi/94/5/JVI.02015–19.atom.

37. Shet A, Ray D, Malavige N, Santosham M, Bar-Zeev N. Differential COVID-19-attributable mortality and BCG vaccine use in countries [Internet]. Infectious Diseases (except HIV/AIDS); 2020 Apr [cited 2020 May 4]. Available from: http://medrxiv.org/lookup/doi/10.1101/2020.04.01.20049478

38. Sala G, Miyakawa T. Association of BCG vaccination policy with prevalence and mortality of COVID-19 [Internet]. Epidemiology; 2020 Apr [cited 2020 May 4]. Available from: http://medrxiv.org/lookup/doi/10.1101/2020.03.30.20048165

39. Hensel J, McGrail DJ, McAndrews KM, Dowlatshahi D, LeBleu VS, Kalluri R. Exercising caution in correlating COVID-19 incidence and mortality rates with BCG vaccination policies due to variable rates of SARS CoV-2 testing [Internet]. Epidemiology; 2020 Apr [cited 2020 May 4]. Available from: http://medrxiv.org/lookup/doi/10.1101/2020.04.08.20056051

40. Szigeti R, Kellermayer D, Kellermayer R. BCG protects against COVID-19? A word of caution [Internet]. Epidemiology; 2020 Apr [cited 2020 May 4]. Available from: http://medrxiv.org/lookup/doi/10.1101/2020.04.09.20056903

41. Franklin R, Young A, Neumann B, Fernandez R, Joannides A, Reyahi A, et al. Homologous protein domains in SARS-CoV-2 and measles, mumps and rubella viruses: preliminary evidence that MMR vaccine might provide protection against COVID-19 [Internet]. Infectious Diseases (except HIV/AIDS); 2020 Apr [cited 2020 May 4]. Available from: http://medrxiv.org/lookup/doi/10.1101/2020.04.10.20053207

42. Jin H, Li Y, Ma Z, Zhang F, Xie Q, Gu D, et al. Effect of chemical adjuvants on DNA vaccination. Vaccine. 2004 Jul;22(21–22):2925–35.

43. Petrova M, Jelev D, Ivanova A, Krastev Z. Isoprinosine Affects Serum Cytokine Levels in Healthy Adults. Journal of Interferon & Cytokine Research. 2010 Apr;30(4):223–8.

44. Villalaín J. Membranotropic Effects of Arbidol, a Broad Anti-Viral Molecule, on Phospholipid Model Membranes. J Phys Chem B. 2010 Jul;114(25):8544–54.

45. Deng L, Li C, Zeng Q, Liu X, Li X, Zhang H, et al. Arbidol combined with LPV/r versus LPV/r alone against Corona Virus Disease 2019: A retrospective cohort study. Journal of Infection. 2020Mar;S0163445320301134.

46. Jeon S, Ko M, Lee J, Choi I, Byun SY, Park S, et al. Identification of antiviral drug candidates against SARS-CoV-2 from FDA-approved drugs [Internet]. Microbiology; 2020 Mar [cited 2020 May 4]. Available from: http://biorxiv.org/lookup/doi/10.1101/2020.03.20.999730

47. Fan H-H, Wang L-Q, Liu W-L, An X-P, Liu Z-D, He X-Q, et al. Repurposing of clinically approved drugs for treatment of coronavirus disease 2019 in a 2019-novel coronavirus-related coronavirus model. Chin Med J. 2020 May 5;133(9):1051–6.

48. Dyall J, Coleman CM, Hart BJ, Venkataraman T, Holbrook MR, Kindrachuk J, et al. Repurposing of Clinically Developed Drugs for Treatment of Middle East Respiratory Syndrome Coronavirus Infection. Antimicrob Agents Chemother. 2014 Aug;58(8):4885–93.

49. Elfiky AA. Ribavirin, Remdesivir, Sofosbuvir, Galidesivir, and Tenofovir against SARS-CoV-2 RNA dependent RNA polymerase (RdRp): A molecular docking study. Life Sciences. 2020Jul;253:117592.

50. Lucas J, Heinlein C, Kim T, Hernandez S, Malik M, True L. The androgen-regulated protease TMPRSS2 activates a proteolytic cascade involving components of the tumor microenvironment and promotes prostate cancer metastasis. Cancer discovery. 2014;4:1310–25.

51. Sanders JM, Sanders ML, Jodlowski TZ, Cutrell JB. Pharmacologic Treatments for Coronavirus Disease 2019 (COVID-19): A Review. JAMA [Internet]. 2020 Apr 13 [cited 2020 May 4]; Available from: https://jamanetwork.com/journals/jama/fullarticle/2764727

52. Sheahan TP, Sims AC, Zhou S, Graham RL, Pruijssers AJ, Agostini ML, et al. An orally bioavailable broad-spectrum antiviral inhibits SARS-CoV-2 in human airway epithelial cell cultures and multiple coronaviruses in mice. Sci Transl Med. 2020 Apr 29;12(541):eabb5883.

